# Type II inflammatory comorbidities in patients with psoriasis: A nationwide study based on Chinese registration

**DOI:** 10.1101/2024.12.18.24319258

**Authors:** Jiaying Zhang, Yanjun Li, Dengli Li, Hongxia Yan, Hong jun Fu, Tianhang Li, Ruoyu Li, Ping Cui, Dongmei Shi

## Abstract

**Purpose:** This study aims to investigate the prevalence and patterns of type II inflammatory disease among adult patients with psoriasis in China, including the associated comorbidities and coexisting conditions.

**Patients and methods:** Data were collected from the register of China National Clinical Center for Skin and Immune Diseases between June 2020 and May 2023. Univariate and a multivariable logistic regression models were used to analyze the factors associated with type II inflammatory disease in patients with psoriasis.

**Results:** A total of 15172 adults with psoriasis were included in this study, and the mean age was 42.46 years and 35.21% were females. Among them, 725 were identified to have type II inflammatory disease, resulting in an overall prevalence of 4.78%. Multivariate logistic regression analysis suggested that the odds ratio (95% confidence interval) for type II inflammatory disease in patients with psoriasis was 1.12 (1.06-1.18) for older age (per 10-year increase), 1.05 (0.89-1.23) for female sex, 1.03 (0.81-1.32) for obesity (BMI≥28kg/m2), 1.32 (1.10-1.58) for smoking, 17.21 (10.46-28.30) for hypertension, 3.14 (2.47-3.99) for history of drug allergy, 1.56 (1.28-1.19) for family history of psoriasis, and 0.60 (0.49-0.73) for severity of the disease (severe vs mild psoriasis).

**Conclusion:** Our results indicate that type II inflammatory diseases in patients with psoriasis is associated with smoking, hypertension, history of drug allergy, and family history of psoriasis. Furthermore, we observed a correlation between the severity of psoriasis and a decreased likelihood of type II inflammatory diseases. This study contribute to valuable real-world data with regard to epidemiological features of type II inflammation among patients with psoriasis in China. It also offer insights into potential risk factors for targeted interventions and personalized treatment strategies in clinical setting.

## Introduction

Psoriasis is a chronic, recurrent, inflammatory and systemic disease that is immune-mediated and influenced by genetic and environmental factors.^1^ Immunological and genetic studies have identified key factors, such as IL-17 and IL-23, in the pathogenesis of psoriasis.^1^ Th1 and Th17 cells are believed to play a crucial role in psoriasis-related cellular immunity and inflammatory response, while Th2 cells play a smaller role.

Type II inflammatory diseases refers to overactive immune called type II immune response, which can trigger various inflammatory reactions in different organs, including skin, respiratory system, and digestive system.^2^ Type II inflammatory diseases include atopic dermatitis (AD), eczema, prurigo nodularis (PN), chronic spontaneous urticaria (CSU), bullous pemphigoid (BP), and scabies.^3^ The type II immune response is typically triggered by the destruction of the skin barrier, involving various resident immune cells such as dendritic cells, macrophages, T cells and innate lymphoid cells (ILCs ).^4^ In type II inflammation, Th2 cells play a significant role in immune response by producing cytokines such as IL-4, IL-5 and IL-13. These cytokines regulate B cells proliferation and differentiation, promote IgE production, and participate in the pathogenesis the type II inflammatory skin diseases.^5^ AD is one of the most common and representative type II inflammatory skin diseases, characterized by persistent itching, recurrent episodes, localized eczema that may exhibit seasonal variations.^6^

Psoriasis and AD are often considered as opposing immune responses. In some patients with psoriasis, the use of TNF inhibitors and IL-17 inhibiting biological agents can trigger the onset of AD, leading to eosinophilia, dry skin, and eczema or dermatitis. ^7–9^ Conversely, the AD-specific biological agent dupilumab may induce psoriasis-like skin lesions.^10^ However, some research revealed that psoriasis can manifest as eczema in the acute phase, while AD can present as psoriasis in the chronic phase.^11^ Nevertheless, studies have shown that some types of AD also involve Th17 .^12^ Tapinarof, a new drug for the treatment of psoriasis, has also been shown to be effective for the treatment of AD.^13^ A study in the United States of America and a study on adolescents have shown that asthma and allergic rhinitis are associated with psoriasis. ^14,15^A study also supports a causal effect between inflammatory bowel disease and psoriasis.^16^ Others found a relationship between psoriasis and prevalence of type II inflammatory diseases such as alopecia areata, vitiligo, BP, and asthma.^17^ However, there is a lack of systemic studies to investigate type II inflammatory diseases among patients with psoriasis.

Both psoriasis and type II inflammatory diseases cause great distress to patients, which not only affects appearance but also affects mental health (Figure 1). Therefore, in this study, by using the real-world clinical data from the national register of psoriasis patients, we sought to explore the epidemiology and associated factors of comorbid type II inflammatory disease among patients with psoriasis.

**Fig 1.**
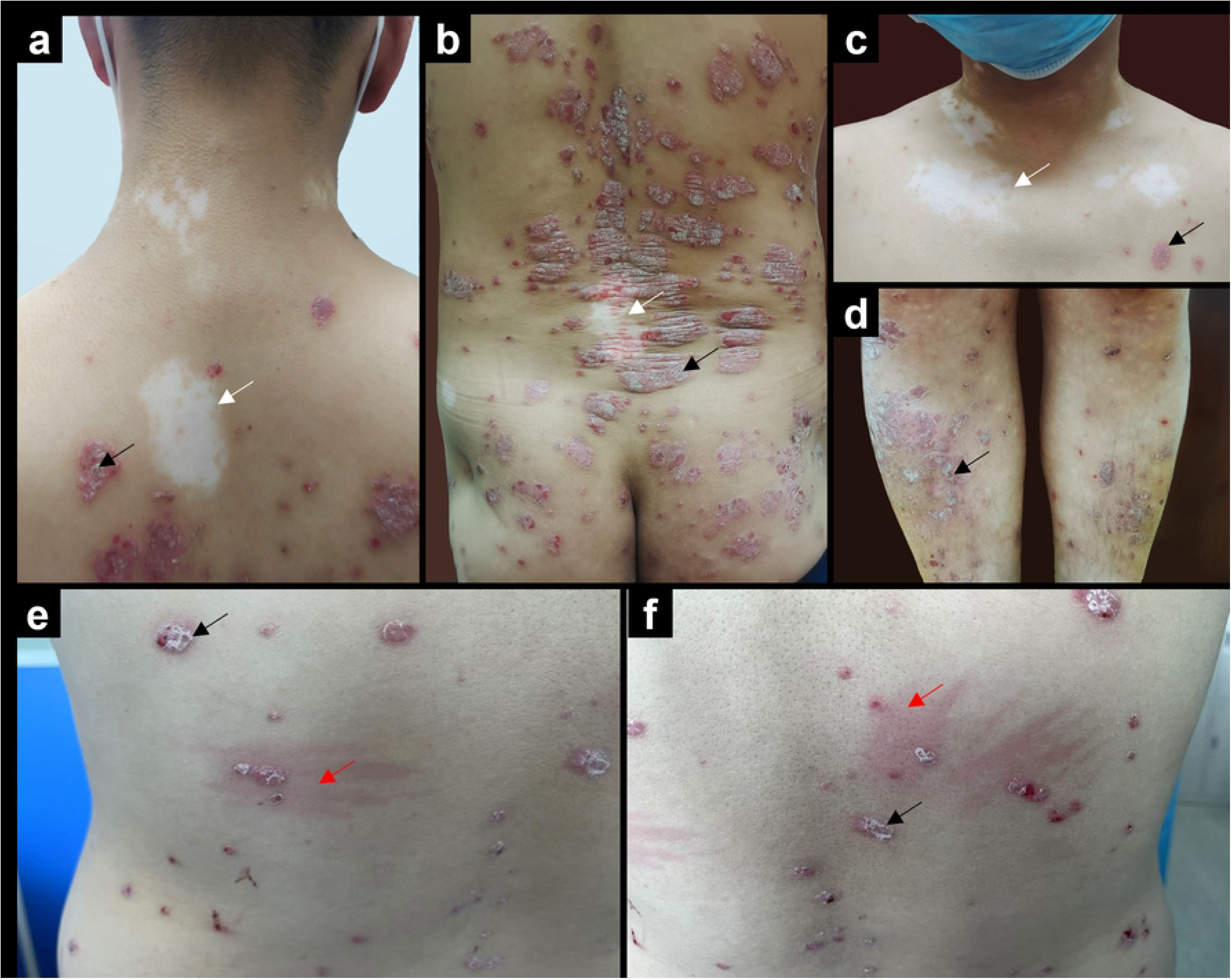
Clinical pictures of patients with psoriasis and Type II inflammatory. (a-d) Psoriasis combined with vitiligo patients with chest, lower limbs and back. (e-f) The back of a patient with psoriasis combined with urticaria. Psoriasis lesions with silvery-white scales and limited papules and plaques are indicated by black arrows. Vitiligo with localised depigmentation indicated by white arrows. Urticaria erythema lesions indicated by red arrows

## Material and methods

### Study design and data source

This was a national register-based study. The study used data derived from the Register of China National Clinical Center for Skin and Immune Diseases, which included a total of 16180 patients who were clinically diagnosed with psoriasis between June 2020 and May 2023 in 215 hospitals across China. Of these, we excluded 1006 patients who were under 18 years of age and 2 patients for missing data on gender, age, BMI, history of psoriasis and type II inflammatory disease. Thus, the analytical sample included 15172 patients with psoriasis. Figure 2 shows flowchart of the study participants. This study was based on participants providing informed consent in electronic written form. Authors had access to information that could identify individual participants during or after data collection.

**Fig 2.**
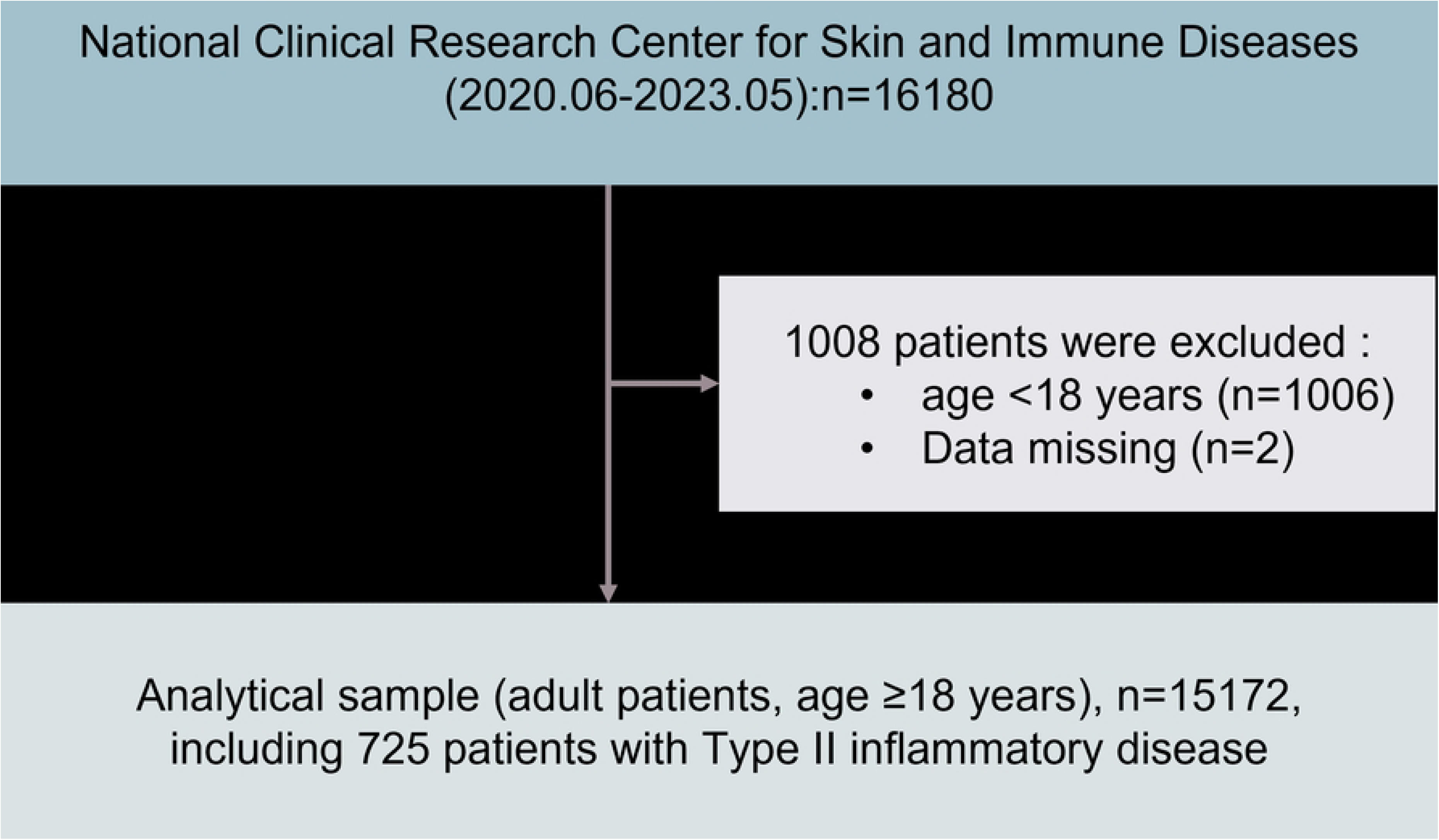
Flowchart of study participants.

### Source databases

The register system included information on age, sex, the first onset age of psoriasis, education (middle school or below, high school, and university or above), height, weight, smoking status, history of drug allergy, and family history of psoriasis.

Body mass index (BMI) was calculated as weight in kg divided by height in meters squared (kg/m^2^), and overweight was defined as BMI of 24-27.9 kg/m^2^ and obesity as BMI ≥28 kg/m^2^.^18^ Early-onset psoriasis (EOP) was defined as psoriasis with age of first onset ≤40 years and late-onset psoriasis (LOP) as age of first onset >40 years.^19^

Information on clinical features of psoriasis recorded in the register included subtype, disease severity, distribution of skin lesion, and etc. In the present study, the types of psoriasis were categorized as plaque psoriasis, guttate psoriasis, erythrodermic psoriasis, pustular psoriasis, and arthritic psoriasis. Severity of psoriasis was determined using the Psoriasis Area Severity Index (PASI),^20^ which assessed factor such as lesion extension, erythema, infiltration, and desquamation using PASI. Psoriasis severity was categorized into mild (PASI score <3), moderate (PASI score 3-9), or severe (PASI score ≥10). The season of exacerbation was self-reported information and classified as spring, summer, autumn, winter, irregular, or seasonal. We defined “predilection season ≥ 2” as irregular.

### Definition of type II inflammatory disease

Type II inflammatory disease in patients with psoriasis is determined by integrating self-reported disease histories. According to the Expert Consensus on the Mechanism and Targeted Therapy of Type II inflammatory Diseases published in the Chinese Medical Journal,^2^ type II inflammatory disease in this study refers to a general term encompassing abnormal type II immune and inflammatory reactions that lead to type II chronic inflammatory diseases. These include conditions such as allergic rhinitis, urticaria, asthma, eczema, vitiligo, AD, chronic obstructive pulmonary disease (COPD), bronchitis, allergic purpura, alopecia areata, allergic dermatitis, chronic sinusitis, ulcerative colitis, scabies, ocular uveitis, BP, PN, food allergy and drug allergy.

### Ethics consideration

This study was approved by the Ethical Committee of the register of China National Clinical Center for Skin and Immune Diseases (2020-255) and Jining No.1 People’s Hospital (2021-084). The electronic informed consent form was obtained from all the patients in the study.

### Statistical analysis

All statistical analyses were performed with IBM SPSS Statistics for Windows, version 26.0 (IBM Incorp, Armonk, NY, USA). Characteristics of study participants are presented as frequencies (%) for categorical variables and means (standard deviation, SD) for continuous variables. The chi-square test was used to compare the patients’ characteristics of by sex and type II inflammatory disease status. Univariate and multivariate logistic regression was used to estimate the odds ratio (OR) and 95% confidence intervals (CIs) of type II inflammatory disease in association with various factors among psoriasis patients. Two-tailed P<0.05 was considered to be statistically significant.

## Results

### Characteristics of study participants

This study enrolled 16,180 patients with psoriasis. After excluding 2 patients with insufficient information and 1006 patients under18 years old, the remaining sample size was 15,172. Among them, 64.76 % (9826/15172) were males 35.21 % (5342/15172) were females, resulting in a male-to-female ratio of 1.84. There were also 4 patients (0.03%) with missing gender information. The average age of the patients was 42.46 years and the average age of onset was 36.25 years.

Male patients with psoriasis, compared to females, exhibited older age, higher rates of overweight or obesity, and higher education levels, and higher rates of smoking (P<0.05). Male patients are also more likely to have hypertension, but less likely to have a history of drug allergy or a family history of psoriasis compared to female patients (P<0.05).

Both male and female patients commonly presented with plaque psoriasis, followed by guttate psoriasis. However, there were notable difference in the following types between gender. Among males, the third most common type was erythrodermic psoriasis, followed by arthropathic psoriasis and pustular psoriasis, while among females, the third most common type was pustular psoriasis, followed by arthropathic psoriasis and erythrodermic psoriasis. There were statistically significant differences between male and female patients in terms of plaque, guttate, pustular, and erythrodermic psoriasis (P<0.001 for plaque, guttate, and pustular; P<0.05 for erythrodermic). There was no significant difference between genders in terms of arthritis psoriasis. Furthermore, the proportion of male patients with severe PASI scores was higher compared to female patients (P< 0.001).

Both male and female patients experienced irregular onset of the condition, with exacerbations occurring more freqently in winter and then in spring. The distribution of skin lesions in both genders predominantly mixed types, indicating the presence of two difference lesion types. Additionally, male patients exhibited a higher proportion of skik lesions on various body areas including the scalp, face, neck, chest, back, upper limbs, lower limbs, hands, feet and genitals (Table 1).

**Table 1.**
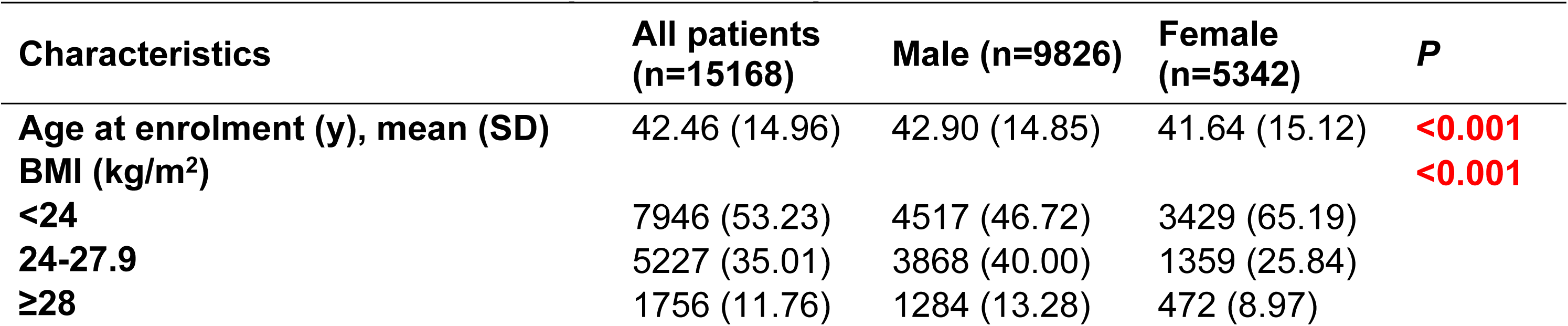
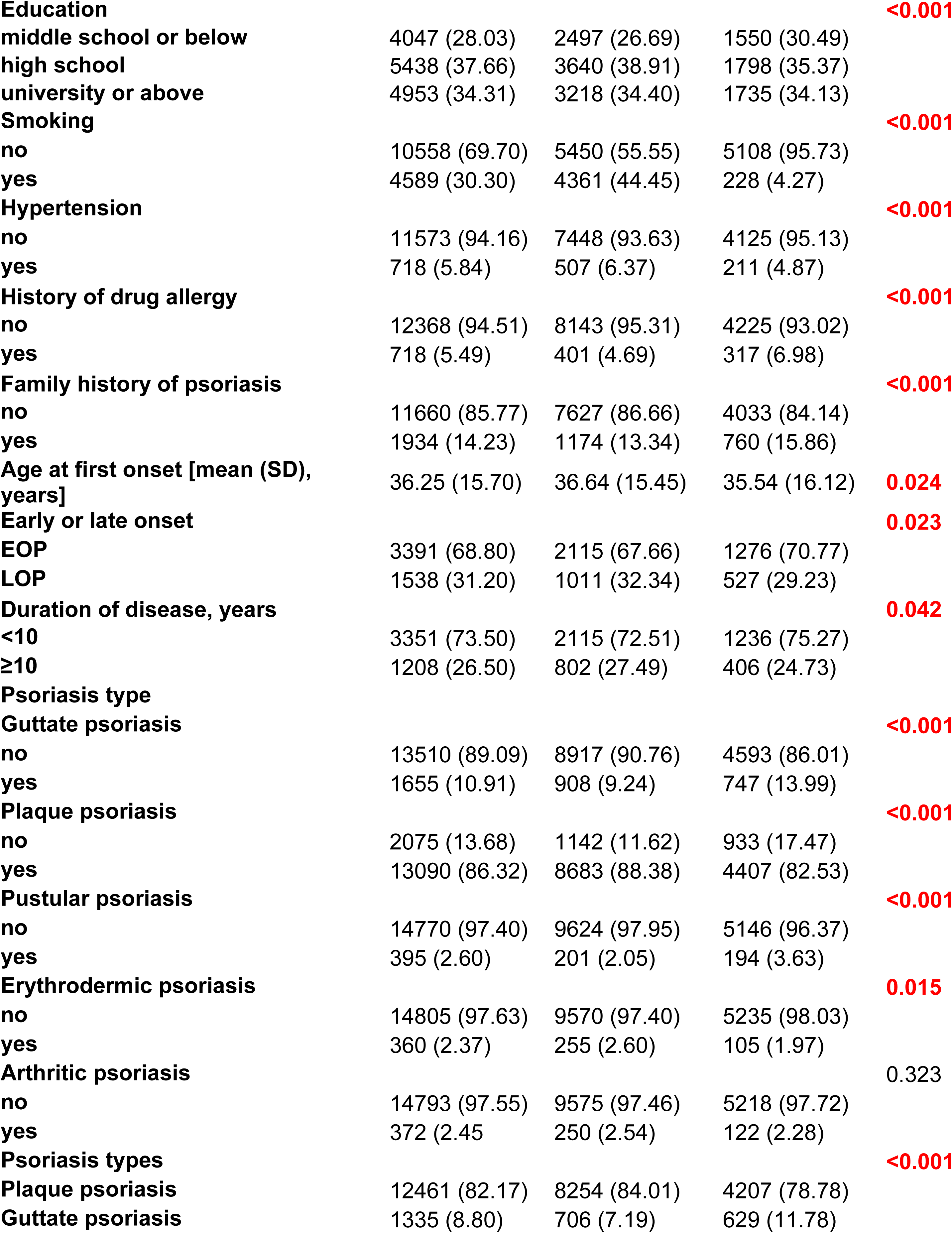
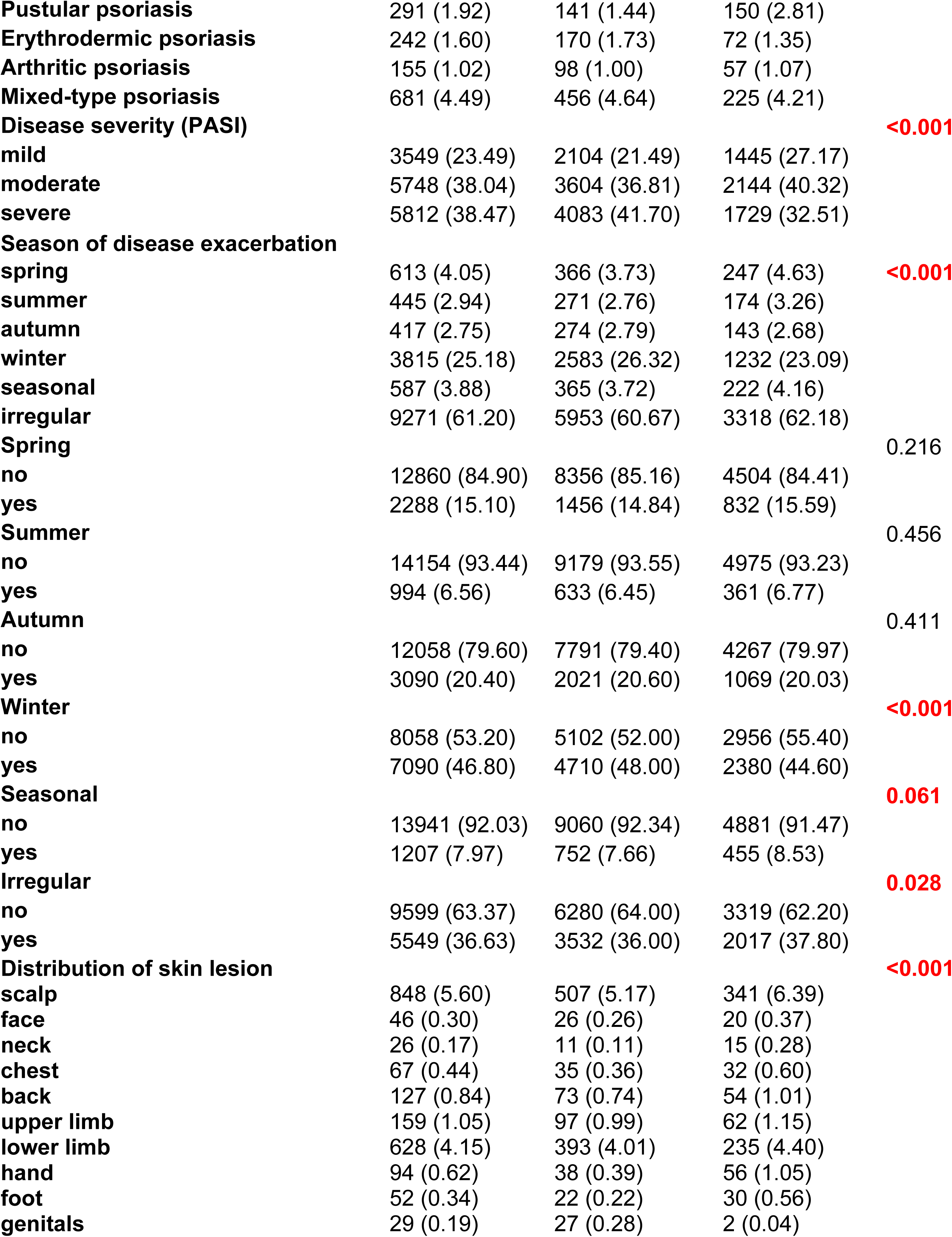
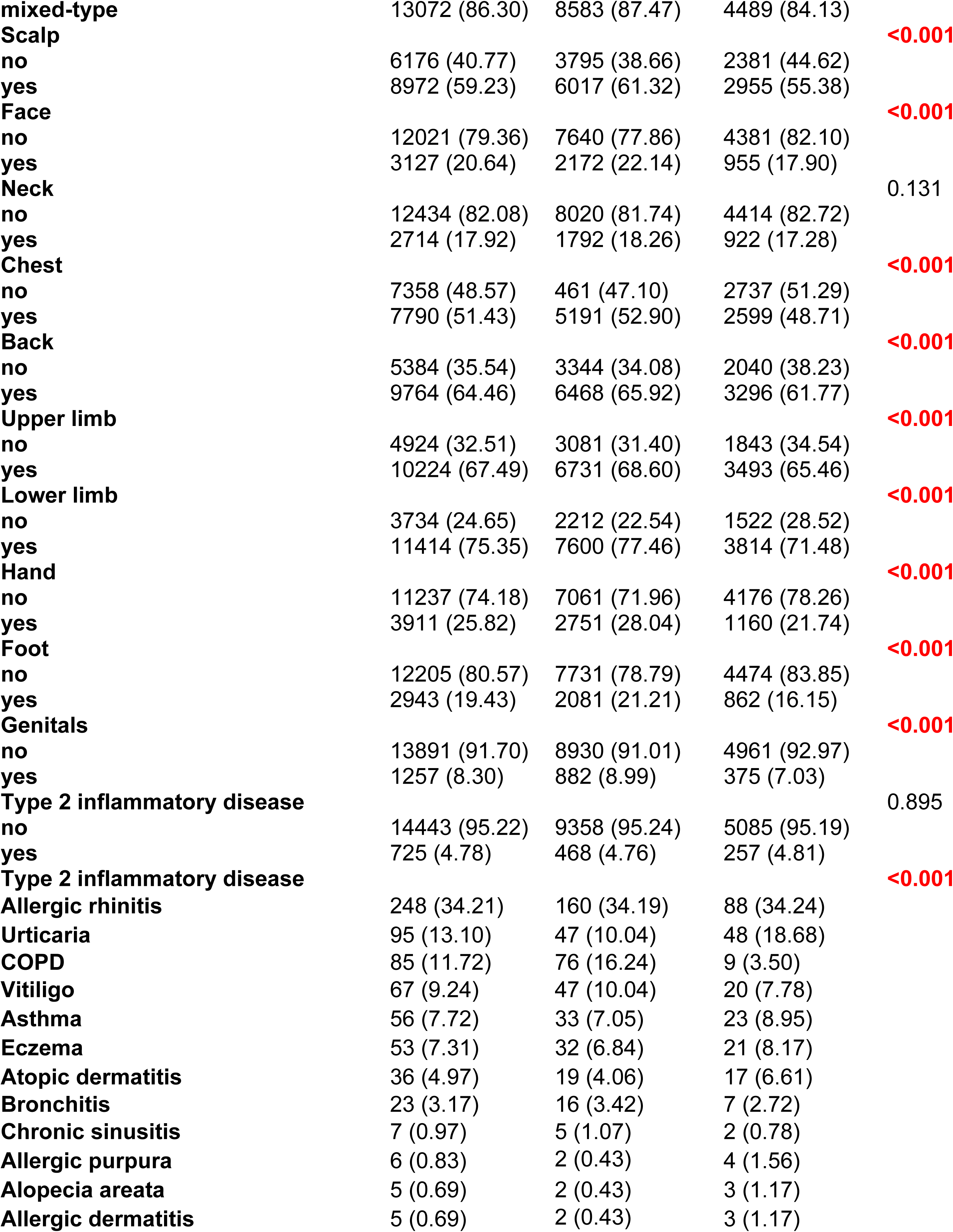
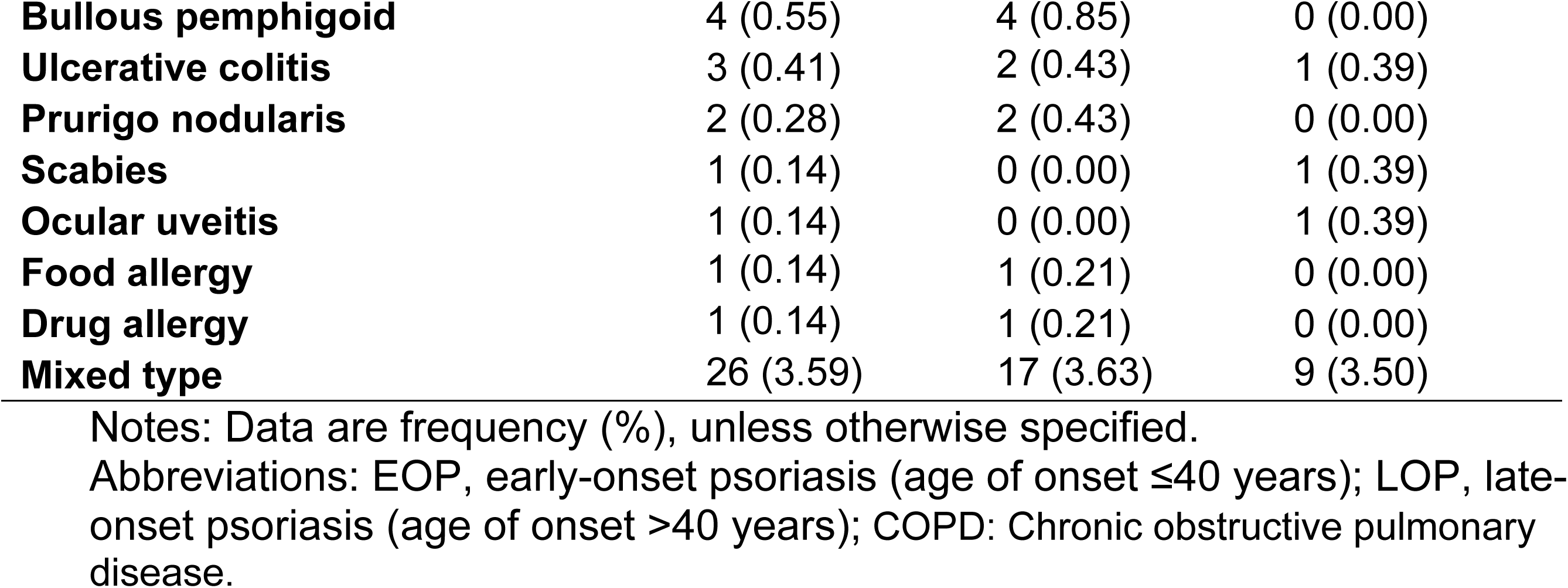
Characteristics of patients with psoriasis.

| Characteristics | All patients (n=15168) | Male (n=9826) | Female (n=5342) | P |
| --- | --- | --- | --- | --- |
| Age at enrolment (y), mean (SD) | 42.46 (14.96) | 42.90 (14.85) | 41.64 (15.12) | **<0.001** |
| BMI (kg/m<sup>2</sup>) |  |  |  | **<0.001** |
| <24 | 7946 (53.23) | 4517 (46.72) | 3429 (65.19) |  |
| 24-27.9 | 5227 (35.01) | 3868 (40.00) | 1359 (25.84) |  |
| ≥28 | 1756 (11.76) | 1284 (13.28) | 472 (8.97) |  |
| **Education** |  |  |  | **<0.001** |
| middle school or below | 4047 (28.03) | 2497 (26.69) | 1550 (30.49) |  |
| high school | 5438 (37.66) | 3640 (38.91) | 1798 (35.37) |  |
| university or above | 4953 (34.31) | 3218 (34.40) | 1735 (34.13) |  |
| **Smoking** |  |  |  | **<0.001** |
| no | 10558 (69.70) | 5450 (55.55) | 5108 (95.73) |  |
| yes | 4589 (30.30) | 4361 (44.45) | 228 (4.27) |  |
| **Hypertension** |  |  |  | **<0.001** |
| no | 11573 (94.16) | 7448 (93.63) | 4125 (95.13) |  |
| yes | 718 (5.84) | 507 (6.37) | 211 (4.87) |  |
| **History of drug allergy** |  |  |  | **<0.001** |
| no | 12368 (94.51) | 8143 (95.31) | 4225 (93.02) |  |
| yes | 718 (5.49) | 401 (4.69) | 317 (6.98) |  |
| **Family history of psoriasis** |  |  |  | **<0.001** |
| no | 11660 (85.77) | 7627 (86.66) | 4033 (84.14) |  |
| yes | 1934 (14.23) | 1174 (13.34) | 760 (15.86) |  |
| **Age at first onset [mean (SD), years]** | 36.25 (15.70) | 36.64 (15.45) | 35.54 (16.12) | **0.024** |
| **Early or late onset** |  |  |  | **0.023** |
| EOP | 3391 (68.80) | 2115 (67.66) | 1276 (70.77) |  |
| LOP | 1538 (31.20) | 1011 (32.34) | 527 (29.23) |  |
| **Duration of disease, years** |  |  |  | **0.042** |
| <10 | 3351 (73.50) | 2115 (72.51) | 1236 (75.27) |  |
| ≥10 | 1208 (26.50) | 802 (27.49) | 406 (24.73) |  |
| **Psoriasis type** |  |  |  |  |
| **Guttate psoriasis** |  |  |  | **<0.001** |
| no | 13510 (89.09) | 8917 (90.76) | 4593 (86.01) |  |
| yes | 1655 (10.91) | 908 (9.24) | 747 (13.99) |  |
| **Plaque psoriasis** |  |  |  | **<0.001** |
| no | 2075 (13.68) | 1142 (11.62) | 933 (17.47) |  |
| yes | 13090 (86.32) | 8683 (88.38) | 4407 (82.53) |  |
| **Pustular psoriasis** |  |  |  | **<0.001** |
| no | 14770 (97.40) | 9624 (97.95) | 5146 (96.37) |  |
| yes | 395 (2.60) | 201 (2.05) | 194 (3.63) |  |
| **Erythrodermic psoriasis** |  |  |  | **0.015** |
| no | 14805 (97.63) | 9570 (97.40) | 5235 (98.03) |  |
| yes | 360 (2.37) | 255 (2.60) | 105 (1.97) |  |
| **Arthritic psoriasis** |  |  |  | 0.323 |
| no | 14793 (97.55) | 9575 (97.46) | 5218 (97.72) |  |
| yes | 372 (2.45) | 250 (2.54) | 122 (2.28) |  |
| **Psoriasis types** |  |  |  | **<0.001** |
| Plaque psoriasis | 12461 (82.17) | 8254 (84.01) | 4207 (78.78) |  |
| Guttate psoriasis | 1335 (8.80) | 706 (7.19) | 629 (11.78) |  |
| **Pustular psoriasis** | 291 (1.92) | 141 (1.44) | 150 (2.81) |  |
| **Erythrodermic psoriasis** | 242 (1.60) | 170 (1.73) | 72 (1.35) |  |
| **Arthritic psoriasis** | 155 (1.02) | 98 (1.00) | 57 (1.07) |  |
| **Mixed-type psoriasis** | 681 (4.49) | 456 (4.64) | 225 (4.21) |  |
| **Disease severity (PASI)** |  |  |  | **<0.001** |
| **mild** | 3549 (23.49) | 2104 (21.49) | 1445 (27.17) |  |
| **moderate** | 5748 (38.04) | 3604 (36.81) | 2144 (40.32) |  |
| **severe** | 5812 (38.47) | 4083 (41.70) | 1729 (32.51) |  |
| **Season of disease exacerbation** |  |  |  |  |
| **spring** | 613 (4.05) | 366 (3.73) | 247 (4.63) | **<0.001** |
| **summer** | 445 (2.94) | 271 (2.76) | 174 (3.26) |  |
| **autumn** | 417 (2.75) | 274 (2.79) | 143 (2.68) |  |
| **winter** | 3815 (25.18) | 2583 (26.32) | 1232 (23.09) |  |
| **seasonal** | 587 (3.88) | 365 (3.72) | 222 (4.16) |  |
| **irregular** | 9271 (61.20) | 5953 (60.67) | 3318 (62.18) |  |
| **Spring** |  |  |  | 0.216 |
| **no** | 12860 (84.90) | 8356 (85.16) | 4504 (84.41) |  |
| **yes** | 2288 (15.10) | 1456 (14.84) | 832 (15.59) |  |
| **Summer** |  |  |  | 0.456 |
| **no** | 14154 (93.44) | 9179 (93.55) | 4975 (93.23) |  |
| **yes** | 994 (6.56) | 633 (6.45) | 361 (6.77) |  |
| **Autumn** |  |  |  | 0.411 |
| **no** | 12058 (79.60) | 7791 (79.40) | 4267 (79.97) |  |
| **yes** | 3090 (20.40) | 2021 (20.60) | 1069 (20.03) |  |
| **Winter** |  |  |  | **<0.001** |
| **no** | 8058 (53.20) | 5102 (52.00) | 2956 (55.40) |  |
| **yes** | 7090 (46.80) | 4710 (48.00) | 2380 (44.60) |  |
| **Seasonal** |  |  |  | **0.061** |
| **no** | 13941 (92.03) | 9060 (92.34) | 4881 (91.47) |  |
| **yes** | 1207 (7.97) | 752 (7.66) | 455 (8.53) |  |
| **Irregular** |  |  |  | **0.028** |
| **no** | 9599 (63.37) | 6280 (64.00) | 3319 (62.20) |  |
| **yes** | 5549 (36.63) | 3532 (36.00) | 2017 (37.80) |  |
| **Distribution of skin lesion** |  |  |  | **<0.001** |
| **scalp** | 848 (5.60) | 507 (5.17) | 341 (6.39) |  |
| **face** | 46 (0.30) | 26 (0.26) | 20 (0.37) |  |
| **neck** | 26 (0.17) | 11 (0.11) | 15 (0.28) |  |
| **chest** | 67 (0.44) | 35 (0.36) | 32 (0.60) |  |
| **back** | 127 (0.84) | 73 (0.74) | 54 (1.01) |  |
| **upper limb** | 159 (1.05) | 97 (0.99) | 62 (1.15) |  |
| **lower limb** | 628 (4.15) | 393 (4.01) | 235 (4.40) |  |
| **hand** | 94 (0.62) | 38 (0.39) | 56 (1.05) |  |
| **foot** | 52 (0.34) | 22 (0.22) | 30 (0.56) |  |
| **genitals** | 29 (0.19) | 27 (0.28) | 2 (0.04) |  |
| **mixed-type** | 13072 (86.30) | 8583 (87.47) | 4489 (84.13) |  |
| **Scalp** |  |  |  | **<0.001** |
| no | 6176 (40.77) | 3795 (38.66) | 2381 (44.62) |  |
| yes | 8972 (59.23) | 6017 (61.32) | 2955 (55.38) |  |
| **Face** |  |  |  | **<0.001** |
| no | 12021 (79.36) | 7640 (77.86) | 4381 (82.10) |  |
| yes | 3127 (20.64) | 2172 (22.14) | 955 (17.90) |  |
| **Neck** |  |  |  | 0.131 |
| no | 12434 (82.08) | 8020 (81.74) | 4414 (82.72) |  |
| yes | 2714 (17.92) | 1792 (18.26) | 922 (17.28) |  |
| **Chest** |  |  |  | **<0.001** |
| no | 7358 (48.57) | 461 (47.10) | 2737 (51.29) |  |
| yes | 7790 (51.43) | 5191 (52.90) | 2599 (48.71) |  |
| **Back** |  |  |  | **<0.001** |
| no | 5384 (35.54) | 3344 (34.08) | 2040 (38.23) |  |
| yes | 9764 (64.46) | 6468 (65.92) | 3296 (61.77) |  |
| **Upper limb** |  |  |  | **<0.001** |
| no | 4924 (32.51) | 3081 (31.40) | 1843 (34.54) |  |
| yes | 10224 (67.49) | 6731 (68.60) | 3493 (65.46) |  |
| **Lower limb** |  |  |  | **<0.001** |
| no | 3734 (24.65) | 2212 (22.54) | 1522 (28.52) |  |
| yes | 11414 (75.35) | 7600 (77.46) | 3814 (71.48) |  |
| **Hand** |  |  |  | **<0.001** |
| no | 11237 (74.18) | 7061 (71.96) | 4176 (78.26) |  |
| yes | 3911 (25.82) | 2751 (28.04) | 1160 (21.74) |  |
| **Foot** |  |  |  | **<0.001** |
| no | 12205 (80.57) | 7731 (78.79) | 4474 (83.85) |  |
| yes | 2943 (19.43) | 2081 (21.21) | 862 (16.15) |  |
| **Genitals** |  |  |  | **<0.001** |
| no | 13891 (91.70) | 8930 (91.01) | 4961 (92.97) |  |
| yes | 1257 (8.30) | 882 (8.99) | 375 (7.03) |  |
| **Type 2 inflammatory disease** |  |  |  | 0.895 |
| no | 14443 (95.22) | 9358 (95.24) | 5085 (95.19) |  |
| yes | 725 (4.78) | 468 (4.76) | 257 (4.81) |  |
| **Type 2 inflammatory disease** |  |  |  | **<0.001** |
| **Allergic rhinitis** | 248 (34.21) | 160 (34.19) | 88 (34.24) |  |
| **Urticaria** | 95 (13.10) | 47 (10.04) | 48 (18.68) |  |
| **COPD** | 85 (11.72) | 76 (16.24) | 9 (3.50) |  |
| **Vitiligo** | 67 (9.24) | 47 (10.04) | 20 (7.78) |  |
| **Asthma** | 56 (7.72) | 33 (7.05) | 23 (8.95) |  |
| **Eczema** | 53 (7.31) | 32 (6.84) | 21 (8.17) |  |
| **Atopic dermatitis** | 36 (4.97) | 19 (4.06) | 17 (6.61) |  |
| **Bronchitis** | 23 (3.17) | 16 (3.42) | 7 (2.72) |  |
| **Chronic sinusitis** | 7 (0.97) | 5 (1.07) | 2 (0.78) |  |
| **Allergic purpura** | 6 (0.83) | 2 (0.43) | 4 (1.56) |  |
| **Alopecia areata** | 5 (0.69) | 2 (0.43) | 3 (1.17) |  |
| **Allergic dermatitis** | 5 (0.69) | 2 (0.43) | 3 (1.17) |  |

|  |  |  |  |
| --- | --- | --- | --- |
| **Bullous pemphigoid** | 4 (0.55) | 4 (0.85) | 0 (0.00) |
| **Ulcerative colitis** | 3 (0.41) | 2 (0.43) | 1 (0.39) |
| **Prurigo nodularis** | 2 (0.28) | 2 (0.43) | 0 (0.00) |
| **Scabies** | 1 (0.14) | 0 (0.00) | 1 (0.39) |
| **Ocular uveitis** | 1 (0.14) | 0 (0.00) | 1 (0.39) |
| **Food allergy** | 1 (0.14) | 1 (0.21) | 0 (0.00) |
| **Drug allergy** | 1 (0.14) | 1 (0.21) | 0 (0.00) |
| **Mixed type** | 26 (3.59) | 17 (3.63) | 9 (3.50) |

Notes: Data are frequency (%), unless otherwise specified.
Abbreviations: EOP, early-onset psoriasis (age of onset $\leq 40$ years); LOP, late-onset psoriasis (age of onset $> 40$ years); COPD: Chronic obstructive pulmonary disease.

### Distribution of type II inflammatory diseases in patients with psoriasis

The overall prevalence of type II inflammatory diseases in psoriasis patients was 4.78 % (725/15172), with the prevalence being 3.084 % (468/15172) in males and 1.69 % (257/15172) in females. The age- and sex-specific prevalence of type II inflammatory diseases in patients with psoriasis is shown in Figure 3. The prevalence of type II inflammatory disease in patients with psoriasis showed two peaks with increasing age. The first peak occurred in the age group of 18-24 years old, with the prevalence being 5.33% (4.71% in males and 6.21% in females). The second peak occurred in patients over 64 years old, with the prevalence being 8.49 % (10.67 % in males and 4.21 % in females). Furthermore, the age-specific prevalence of cardiovascular disease was higher in males compared to women.

**Fig 3.**
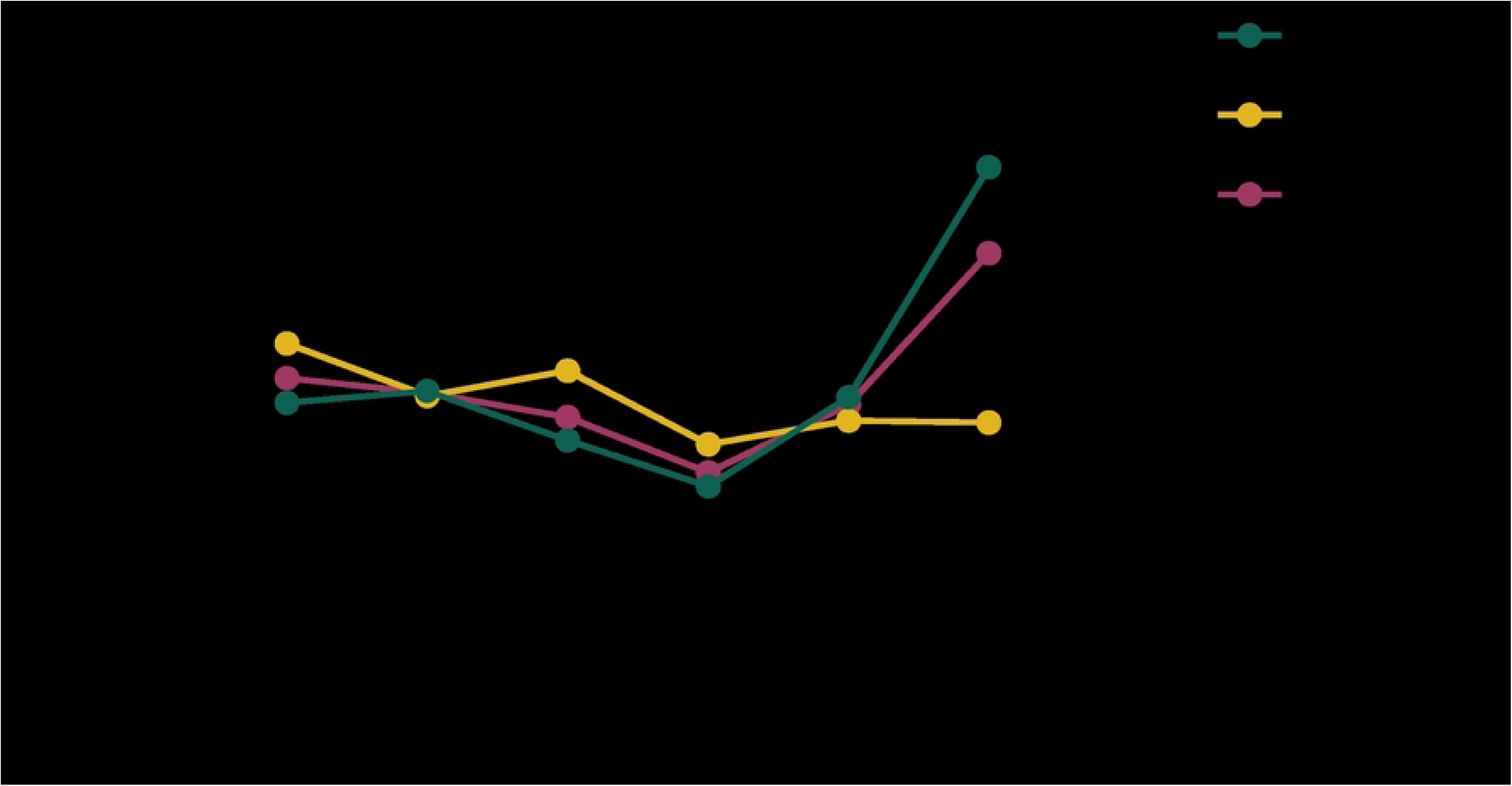
The age- and sex-specific prevalence (per 100 populations) of comorbid type II inflammatory disease in patients with psoriasis.

### Factors associated with type II inflammatory disease in patients with psoriasis

The multivariate logistic regression analysis identified several factors associated with type II inflammatory diseases in psoriasis patients. These factors includeolder age, higher education, smoking, longer duration of psoriasis, early onset and late onset. Additionally, a history of hypertension, drug allergy, or a family history of psoriasis were significantly associated with type II inflammatory diseases.

Furthermore, moderate and severe (compared to mild) psoriasis were linked to type II inflammatory diseases. A dose-response relationship was observed between disease severity and type II inflammatory disease in patients with psoriasis (p for linear trend < 0.001).

Regarding seasonal aggravation, psoriasis exacerbation in winter showed a significantly association with type II inflammatory diseases. However, there was no statistically significant correlation between aggravation in other seasons or irregular aggravation and type II inflammatory diseases (Table 2). The distribution of skin lesions did not show a significant association with type II inflammatory diseases (Table 2).

**Table 2.** Aassociations of comorbid Type II inflammation with demographic, lifestyle, and clinical factors in patients with psoriasis.

| Factors | Type II inflammation, n (%) |  | Odds ratio (95% confidence interval) |  |
| --- | --- | --- | --- | --- |
|  | No, n=14447 | Yes, n=725 | Crude model | Adjusted model<sup>a</sup> |
| **Age at enrolment (per 10-year increment)** | 42.40 (14.82) | 43.76 (17.45) | **1.06 (1.01, 1.12)** | **1.12 (1.06, 1.18)** |
| **Sex** |  |  |  |  |
| male | 9358 (64.79) | 468 (64.55) | 1.00 (ref.) | 1.00 (ref.) |
| female | 5085 (35.21) | 257 (35.45) | 1.01 (0.87, 1.18) | 1.05 (0.89, 1.23) |
| **Education** |  |  |  |  |
| middle school or below | 3886 (28.27) | 163 (23.45) | 1.00 (ref.) | 1.00 (ref.) |
| high school | 5211 (37.90) | 228 (32.81) | 1.04 (0.85, 1.28) | 1.14 (0.92, 1.40) |
| university or above | 4651 (33.83) | 304 (43.74) | **1.56 (1.28, 1.89)** | **1.84 (1.49, 2.28)** |
| **BMI (kg/m<sup>2</sup>)** |  |  |  |  |
| <24 | 7546 (53.10) | 400 (55.63) | 1.00 (ref.) | 1.00 (ref.) |
| 24-27.9 | 4997 (35.17) | 230 (31.99) | 0.87 (0.74, 1.03) | 0.87 (0.73, 1.03) |
| ≥28 | 1667 (11.73) | 89 (12.38) | 1.01 (0.80, 1.28) | 1.03 (0.81, 1.32) |
| **Smoking** |  |  |  |  |
| no | 10089 (69.89) | 475 (66.25) | 1.00 (ref.) | 1.00 (ref.) |
| yes | 4347 (30.11) | 242 (33.75) | **1.18 (1.01, 1.39)** | **1.32 (1.10, 1.58)** |
| Hypertension |  |  |  |  |
| no | 11548 (94.61) | 28 (31.82) | 1.00 (ref.) | 1.00 (ref.) |
| yes | 658 (5.39) | 60 (68.18) | **37.61 (23.85, 59.30)** | **17.21 (10.46, 28.30)** |
| History of drug allergy |  |  |  |  |
| no | 11838 (94.96) | 534 (85.44) | 1.00 (ref.) | 1.00 (ref.) |
| yes | 628 (5.04) | 91 (14.56) | **3.21 (2.54, 4.07)** | **3.14 (2.47, 3.99)** |
| Family history of psoriasis |  |  |  |  |
| no | 11124 (86.07) | 539 (79.97) | 1.00 (ref.) | 1.00 (ref.) |
| yes | 1800 (13.93) | 135 (20.03) | **1.55 (1.27, 1.88)** | **1.56 (1.28, 1.91)** |
| Early or late onset |  |  |  |  |
| EOP<sup>a</sup> | 2521 (65.29) | 501 (71.57) | 1.00 (ref.) | 1.00 (ref.) |
| LOP<sup>b</sup> | 1340 (34.71) | 199 (28.43) | **0.75 (0.63, 0.89)** | **0.37 (0.29, 0.48)** |
| Duration of disease |  |  |  |  |
| <10 years | 2941 (76.17) | 412 (58.86) | 1.00 (ref.) | 1.00 (ref.) |
| ≥10 years | 920 (23.83) | 288 (41.14) | **2.24 (1.89, 2.64)** | **2.13 (1.79, 2.54)** |
| Psoriasis type |  |  |  |  |
| Plaque psoriasis |  |  |  |  |
| no | 1974 (13.66) | 102 (14.07) | 1.00 (ref.) | 1.00 (ref.) |
| yes | 12472 (86.34) | 623 (85.93) | 0.97 (0.78, 1.20) | 0.93 (0.75, 1.16) |
| Guttate psoriasis |  |  |  |  |
| no | 12865 (89.06) | 649 (89.52) | 1.00 (ref.) | 1.00 (ref.) |
| yes | 1581 (10.94) | 76 (10.48) | 0.95 (0.75, 1.22) | 1.00 (0.78, 1.28) |
| Pustular psoriasis |  |  |  |  |
| no | 14071 (97.40) | 705 (97.24) | 1.00 (ref.) | 1.00 (ref.) |
| yes | 375 (2.60) | 20 (2.76) | 1.06 (0.68, 1.68) | 1.04 (0.64, 1.68) |
| Erythrodermic psoriasis |  |  |  |  |
| no | 14100 (97.60) | 711 (98.07) | 1.00 (ref.) | 1.00 (ref.) |
| yes | 346 (2.40) | 14 (1.93) | 0.80 (0.47, 1.38) | 0.76 (0.42, 1.36) |
| Arthritic psoriasis |  |  |  |  |
| no | 14098 (97.59) | 701 (96.69) | 1.00 (ref.) | 1.00 (ref.) |
| yes | 348 (2.41) | 24 (3.31) | 1.39 (0.91, 2.11) | 1.37 (0.90, 2.10) |
| Psoriasis type |  |  |  |  |
| Plaque psoriasis | 11868 (82.15) | 597 (82.34) | 1.00 (ref.) | 1.00 (ref.) |
| Guttate psoriasis | 1272 (8.81) | 64 (8.83) | 1.00 (0.77, 1.30) | 1.05 (0.80, 1.37) |
| Pustular psoriasis | 277 (1.92) | 14 (1.93) | 1.01 (0.58, 1.73) | 1.09 (0.63, 1.89) |
| Erythrodermic psoriasis | 231 (1.60) | 11 (1.52) | 0.95 (0.51, 1.74) | 0.96 (0.50, 1.82) |
| Arthritic psoriasis | 147 (1.02) | 8 (1.10) | 1.08 (0.53, 2.21) | 1.08 (0.52, 2.21) |
| Mixed | 651 (4.51) | 31 (4.28) | 0.95 (0.65, 1.37) | 0.92 (0.63, 1.35) |
| Disease severity (PASI) |  |  |  |  |
| mild | 3331 (23.12) | 221 (31.35) | 1.00 (ref.) | 1.00 (ref.) |
| moderate | 5490 (38.10) | 259 (36.74) | **0.71 (0.59, 0.86)** | **0.72 (0.60, 0.88)** |
| severe | 5589 (38.79) | 225 (31.91) | **0.61 (0.50, 0.73)** | **0.60 (0.49, 0.73)** |
| P-trend |  |  | **<0.001** |  |
| Season of disease exacerbation |  |  |  |  |
| Irregular | 8852 (61.31) | 423 (59.00) | 1.00 (ref.) | 1.00 (ref.) |
| Spring | 591 (4.09) | 22 (3.07) | 0.78 (0.50, 1.21) | 0.76 (0.49, 1.20) |
| Summer | 426 (2.95) | 19 (2.65) | 0.93 (0.58, 1.49) | 0.97 (0.60, 1.55) |
| Autumn | 399 (2.76) | 18 (2.51) | 0.94 (0.58, 1.53) | 0.90 (0.56, 1.46) |
| Winter | 3600 (24.94) | 217 (30.26) | **1.26 (1.07, 1.49)** | **1.25 (1.05, 1.48)** |
| Seasonal | 569 (3.94) | 18 (2.51) | 0.66 (0.41, 1.07) | 0.67 (0.41, 1.08) |
| Spring |  |  |  |  |
| no | 12258 (84.91) | 608 (84.80) | 1.00 (ref.) | 1.00 (ref.) |
| yes | 2179 (15.09) | 109 (15.20) | 1.01 (0.82, 1.24) | 1.02 (0.80, 1.28) |
| Summer |  |  |  |  |
| no | 13481 (93.38) | 679 (94.70) | 1.00 (ref.) | 1.00 (ref.) |
| yes | 956 (6.62) | 38 (5.30) | 0.79 (0.57, 1.10) | 0.80 (0.56, 1.15) |
| Autumn |  |  |  |  |
| no | 11479 (79.51) | 584 (81.45) | 1.00 (ref.) | 1.00 (ref.) |
| yes | 2958 (20.49) | 133 (18.55) | 0.88 (0.73, 1.07) | **0.81 (0.66, 0.99)** |
| Winter |  |  |  |  |
| no | 7711 (53.41) | 349 (48.68) | 1.00 (ref.) | 1.00 (ref.) |
| yes | 6726 (46.59) | 368 (51.32) | **1.21 (1.04, 1.41)** | 1.16 (0.92, 1.47) |
| Seasonal |  |  |  |  |
| no | 13279 (91.98) | 667 (93.03) | 1.00 (ref.) | 1.00 (ref.) |
| yes | 1158 (8.02) | 50 (6.97) | 0.86 (0.64, 1.15) | 0.84 (0.62, 1.15) |
| Irregular |  |  |  |  |
| no | 9132 (63.25) | 470 (65.55) | 1.00 (ref.) | 1.00 (ref.) |
| yes | 5305 (36.75) | 247 (34.45) | 0.91 (0.77, 1.06) | 0.95 (0.73, 1.24) |
| Distribution of skin lesion |  |  |  |  |
| mixed-type | 12452 (86.25) | 624 (87.03) | 1.00 (ref.) | 1.00 (ref.) |
| scalp | 798 (5.53) | 50 (6.97) | 1.25 (0.93, 1.68) | 1.26 (0.93, 1.70) |
| face | 45 (0.31) | 2 (0.28) | 0.89 (0.22, 3.66) | 0.42 (0.06, 3.06) |
| neck | 26 (0.18) | 0 (0.00) | - | - |
| chest | 67 (0.46) | 0 (0.00) | - | - |
| back | 125 (0.87) | 3 (0.42) | 0.48 (0.15, 1.51) | 0.32 (0.08, 1.29) |
| upper limb | 155 (1.07) | 4 (0.56) | 0.52 (0.19, 1.39) | 0.50 (0.18, 1.35) |
| lower limb | 601 (4.16) | 27 (3.77) | 0.90 (0.61, 1.33) | 0.90 (0.61, 1.34) |
| hand | 91 (0.63) | 3 (0.42) | 0.66 (0.21, 2.08) | 0.69 (0.22, 2.19) |
| foot | 49 (0.34) | 3 (0.42) | 1.22 (0.38, 3.93) | 1.26 (0.39, 4.08) |
| genitals | 28 (0.19) | 1 (0.14) | 0.71 (0.10, 5.25) | 0.70 (0.10, 5.18) |

Notes: <sup>a</sup>The model was adjusted for age at enrolment, sex, and education. Abbreviations: EOP, early-onset psoriasis (age of onset ≤40 years); LOP, late-onset psoriasis (age of onset >40 years).

## Discussion

This study utilized real-world clinical data to investigate the prevalence and distribution of type II inflammatory diseases among Chinese adult patients with psoriasis. Additionally, this aimed to analyze demographic, lifestyle, and clinical factors associated with comorbid type II inflammatory diseases in the psoriasis patients. We identified several factors associated with type II inflammatory diseases in patients with psoriasis, including older age, high education level, smoking, hypertension, drug allergy history, family history of psoriasis, early onset and delayed onset, course of disease, and severity of psoriasis. These data provide insights into the epidemiological features of type II inflammatory diseases in Chinese adult patients with psoriasis.

Our study revealed a relative high proportion of comorbid type II inflammatory diseases in psoriasis patients, including allergic rhinitis, COPD, bronchial asthma, and AD among. The prevalence rates of these diseases in the general population in China are approximately 5%-13% for COPD,^21^ over 20% for allergic rhinitis (AR),^22^ around 4.2% for bronchial asthma,^23^ and 2.46% for AD.^24^ However, in our psoriasis patient sample, 4.78% had type II inflammatory related with COPD, allergic rhinitis, bronchial asthma and AD accounted for 0.59%, 1.75%, 0.42%, 0.28%, respectively. These findings suggests a lower prevalence of type II inflammatory diseases in the psoriasis population compared to the general population, which aligns with previous studies indicating that atopy may provide protection against autoimmune diseases like psoriasis.^25^ It’s important to consider that the prevalence rates can vary depending on demographic characteristics of the study populations. Furthermore, the relationship between psoriasis and type II inflammatory diseases is complex and further studies are needed to fully understand the mechanisms and associations between these conditions.

We also observed significant gender differences in specific type II inflammatory diseases among our psoriasis patient sample. The type II inflammatory diseases in our study were COPD, bronchitis, chronic sinusitis, asthma, allergic rhinitis, AD, urticaria, BP, Eczema, PN, Scabies, Vitiligo, alopecia areata, allergic dermatitis, allergic purpura, ocular uveitis, ulcerative colitis, Food allergy, drug allergy and mixed type. Figure 4 illustrates the proportions of these diseases in male, female and overall. There are significant differences in specific diseases between genders. The top three of type II inflammatory diseases among male psoriasis patients are allergic rhinitis, COPD, and urticaria, while in women, they are allergic rhinitis, urticaria, and asthma. The prevalence of COPD has shown an increasing among women in recent years, but it still remains higher in men than that in women.^26^ This can be attributed to higher number of male patients who smoke in our psoriasis patient sample. Additionally, studies have shown a higher prevalence of urticaria is in adult women compared to men,^27^ which is consistent with our findings.

**Fig 4.**
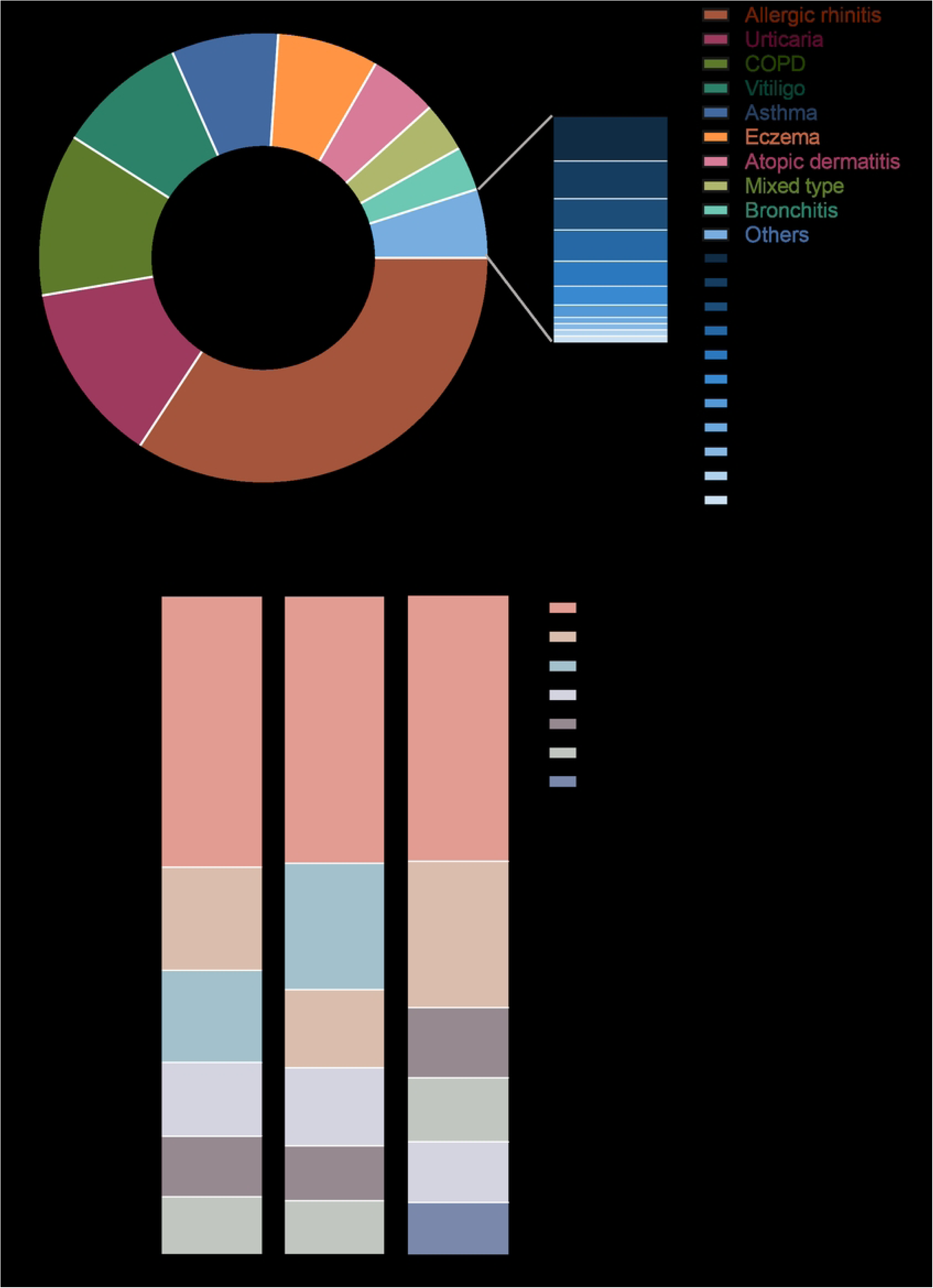
Gender differences in the types of type II inflammatory diseases. (a) Quantitative ranking of all type 2 inflammatory disease types. (b)Top six type of gender differences in type II inflammation types. COPD: chronic obstructive pulmonary disease

Our results show an average age of onset at 33 years old with no gender difference in the prevalence of psoriasis. The highest incidence was in the youth and the elderly, showing a bimodal trend, with women experiencing earlier onset.^28^ Similarly, a bimodal trend was observed in the prevalence of type II inflammatory diseases among psoriasis patients based on age. The first peak occurred in the 18-24 age group, followed by a gradual decrease until 45-54 age group. Thereafter, the prevalence rate sharply increased with age, reaching a second peak in the >64 age group (Figure4). The age of onset varies for different type II inflammatory diseases type II inflammatory. In our sample, allergic rhinitis, urticaria and COPD were the most common type II inflammation diseases. The prevalence of allergic rhinitis peaked in the group of 20-29 years of age and then gradually decreased with age.^29^ A South Korean study demonstrated that CSU is higher incidence in patients with a childhood family history of allergies.^30^ A Canadian study showed that an average age of COPD was 64.7 years old.^31^ Our patients were mostly the outpatients, and the severity of the disease is more likely to be mild (vs. hospitalized patients), and the age may be slightly younger. At present, there is no relevant study on the overall age of onset of type II inflammatory diseases, but the results of the above literature are consistent with our results.

Our study found association between education, smoking, and hypertension with type II inflammatory diseases in psoriasis patients. Higher education levels may contribute to increased mental stress, potentially exacerbating inflammatory conditions. Smoking and hypertension were associated with psoriasis accompanied by type II inflammatory diseases. Smoking was associated with an increased risk of psoriasis, particularly among current smokers. Studies have shown that the risk of psoriasis gradually decreased over time after smoking cessation, highlighting the potential benefits of smoking cessation programs for mitigating the adverse effects on psoriasis.^32^ Hypertension is a common complication of psoriasis, with a higher prevalence being observed in patients with psoriatic arthritis compared to those with psoriasis alone.^33^

Interestingly, we observed a negative correlation between the severity of psoriasis (PASI) and the presence of type II inflammatory diseases. This may be due to the fact that psoriasis is driven by TNF-α and IL-23/Th-17-related pathways,^34^ while type II inflammatory diseases are mainly mediated by Th2-mediated inflammatory responses. Treatment with biological agents for psoriasis patient can induce type II inflammatory diseases such as AD and inflammatory bowel disease.^35,36^ Conversely, the use of biological agents in AD has been reported to induce psoriasis.^37,38^ These diseases can coexist or even overlap, especially with targeted inhibitory effects on Th1 and Th2 immunity. Therefore, severe psoriasis is less likely to be complicated with type II inflammatory diseases, which aligns with clinical observations.

Our study revealed an average psoriasis onset age of 33, with no gender disparity in prevalence. Psoriasis peaks in youth and the elderly, with a similar trend for type II inflammatory diseases. Notably, education, smoking, and hypertension correlate with these diseases in psoriasis patients. Intriguingly, we found a negative correlation between psoriasis severity (PASI) and type II inflammatory diseases presence. This may stem from psoriasis being TNF-α and IL-23/Th-17-related, while type II inflammatory diseases are Th2-mediate.

### Limitation

Some limitations of our study deserve mentioning. Firstly, this study is a retrospective study by nature, meaning that we relied solely on medical records to obtains information. We did not have access to effective tools for physical examination in identification of type II inflammatory diseases. Additionally, our study sample primarily consisted of outpatients, which may introduce a bias towards younger and milder patients. Furthermore, since our data was based on medical records, there is a possibility of incomplete or missing information. It is important to note that the association between psoriasis and type II inflammatory diseases may be underestimated in our study. Therefore, a well-designed large sample study is needed to provide a basis for future research.

## Conclusion

This study represents the first multicenter cross-sectional study utilizing a real-world national database of patients with psoriasis in China. Our findings indicate that the prevalence of type II inflammatory diseases increases tends to increase as the duration of psoriasis progress. Therefore, long-term follow-up of patients with psoriasis is crucial for early detection, intervention and treatment of type II inflammatory diseases. However, it remains a challenge to ascertain whether psoriasis is an independent risk factor or shares a common pathogenesis with type II inflammatory diseases. Exploring more effective treatment methods based on our understanding of psoriasis combined with type II inflammatory diseases, is necessary to reduce complications. Further basic research can provide additional evidence and verify the association between the two diseases.

## Disclosure

The authors report no conflicts of interest in this work.

## Data Availability

Data cannot be shared publicly because of [Privacy or confidentiality]. Data are available from the National Clinical Research Centre for Skin and Immune Diseases Institutional Data Access / Ethics Committee for researchers who meet the criteria for access to confidential data.

